# Brain-Body Signatures of Trait Anxiety Uniquely Characterized by Functional Dynamics, Allostatic Load and Gene Expression

**DOI:** 10.1101/2025.10.26.25338821

**Authors:** Jungyoun Janice Min, Jingxuan Bao, Jae Young Baik, Junhao Wen, Yize Zhao, Paul M. Thompson, Li Shen, Duy Duong-Tran

## Abstract

Trait anxiety is an individual disposition marked by heightened anticipation of potential threats under uncertainty. It has been associated with allostatic load, the cumulative physiological cost of chronic stress, suggesting that enduring anxiety vulnerability emerges from brain–body interactions across multiple scales. Yet these domains have largely been examined separately and the temporal dynamics of brain activity remain underexplored. Using data from healthy young adults (LEMON cohort, N = 120), we implemented a graph-attention framework integrating low-frequency (slow-4 and slow-5) fMRI dynamics, structural connectivity and systemic biomarkers through cross-modal attention to predict individual trait anxiety outcome (The State-Trait Anxiety Inventory). Temporal modeling significantly enhanced prediction compared with static or amplitude-based features, highlighting the importance of time-resolved neural information. Model-derived importance mapping identified the limbic and visual systems as core predictive networks. Dynamic functional connectivity revealed that higher trait anxiety was associated with longer occupancy of states marked by strong limbic–default-mode–frontoparietal coupling and shorter occupancy of visually decoupled states. Metabolic and immune markers further contributed to prediction and transcriptomic enrichment linked these networks to neurodevelopmental and synaptic signaling pathways. Together, these findings delineate a temporally dynamic brain and body architecture underlying stable anxiety vulnerability.

## 1 Introduction

Anxiety disorders are among the most prevalent mental illnesses worldwide, affecting approximately 4% of the global population and steadily increasing over the past two decades [1, 2]. A key vulnerability factor for these disorders is trait anxiety [3], a stable personality disposition that shapes one’s anticipatory responses to potential threats under uncertainty [4–7]. Unlike fear, which arises in response to immediate and identifiable danger, anxiety is characterized by sustained apprehension in unpredictable contexts [8]. Given its pervasive impact and rising prevalence, elucidating the biological substrates of trait anxiety is essential for understanding the mechanisms that give rise to anxiety predisposition before clinical symptoms emerge.

Trait anxiety is tightly coupled with systemic physiological burden of chronic stress, often conceptualized as allostatic load. Under stress, activation of the sympathetic nervous system and the hypothalamic–pituitary–adrenal axis mobilizes energy resources through metabolic and neurobiological changes [9]. This adaptive process, termed allostasis, maintains short-term stability [10], but repeated activation accumulates into allostatic load, a physiological burden of chronic stress [11]. Chronic allostatic load manifests as systemic inflammation [12], metabolic dysregulation [13] and cardiovascular strain [14]. Consistent with these systemic alterations, elevated allostatic load has been reported in patients with generalized anxiety disorder and panic disorder [15], underscoring its clinical relevance. Together, these alterations compromise interoceptive and regulatory balance [16], creating a systemic physiological milieu that heightens vulnerability to anxiety.

Prolonged allostatic load, arising from chronic stress exposure, drives structural and functional reorganization of brain circuits in both animal models and human. Associations have been reported between prolonged stress and structural alterations in stress-sensitive regions such as the hippocampus, amygdala and prefrontal cortex [17, 18], poorer structural integrity within limbic pathways in humans [19] and dysregulation of neurotransmitter signaling in rodent models [20]. Repeated stress exposure induces excessive neurotransmitter release and disrupts calcium homeostasis, leading to long-term alterations in neural activity patterns in rodents [21, 22]. These neurobiological changes represent core underpinnings linking allostatic load to anxiety, consistently supported by neuroimaging evidence.

Resting-state fMRI studies of anxiety have primarily focused on either static functional connectivity or amplitude-based measures. While these approaches have revealed abnormalities across large-scale systems, including the limbic, executive control, default mode and sensory networks, their results have been heterogeneous across studies [23–26]. Amplitude-based indices such as the amplitude of low-frequency fluctuations (ALFF), which quantify the power of spontaneous BOLD oscillations in the 0.01–0.1 Hz range, have reported lower frontal ALFF [27, 28] and frequency-specific alterations in the slow-4 and slow-5 bands that differentiate anxious individuals from controls [29]. Complementary diffusion MRI findings further suggest lower white-matter integrity in individuals with high trait anxiety [30]. Taken together, these results indicate that anxiety involves widespread disturbances across both functional and structural networks, in which many of which overlap with circuits engaged in stress regulation and allostatic load, yet these domains have rarely been examined in an integrated framework.

Despite these advances, several important limitations remain in prior research. First, studies of anxiety have largely examined brain network properties and systemic physiology in isolation, with limited attempts to integrate across domains. Second, conventional rs-fMRI methods fail to capture temporal dynamics. Static functional connectivity summarizes brain signals into time-averaged correlations and ALFF reduces them to frequency-specific amplitude. Both approaches thus may overlook fluctuations across time that may be critical for anxiety. Because allostatic load is associated with alterations in neurotransmitter signaling, such effects are likely to manifest as disruptions in temporal dynamics of brain activity, underscoring the need for analytic strategies that preserve time resolution. Third, although genetic influences on anxiety and brain organization are well established [31–36]whether spatial patterns of gene expression align with neural markers were rarely examined in anxiety context. Exploratory efforts are therefore warranted to probe this potential link. Addressing these gaps requires a multilayered approach spanning neural dynamics, structural connectivity, systemic physiology and transcriptomic architecture.

To bridge the gap between neural and physiological perspectives on anxiety, we investigated how temporal brain dynamics and systemic allostatic markers jointly explain individual variability in trait anxiety (**Figure 1a**). Focusing on early adulthood, a developmental window when anxiety vulnerability peaks [2], we examined how temporal brain dynamics and systemic physiology together account for individual differences in trait anxiety. We first tested whether modeling the temporal evolution of low-frequency brain activity improves prediction beyond static or amplitude-based features (**Figure 1b**). By analyzing attention values from the graph attention network, we identified the key neural circuits that drove model predictions of trait anxiety. To further interpret these effects, we characterized recurrent dynamic functional connectivity states and assessed how their dwell patterns relate to anxiety. We then evaluated whether integrating physiological markers enhances predictive stability compared with neural features alone. Finally, we used attention-based interpretation and transcriptomic enrichment analyses to identify the underlying molecular pathways. Together, this multiscale framework connects neural dynamics, systemic physiology and gene expression, offering a unified account of the biological organization underlying anxiety vulnerability.

**Figure 1.**
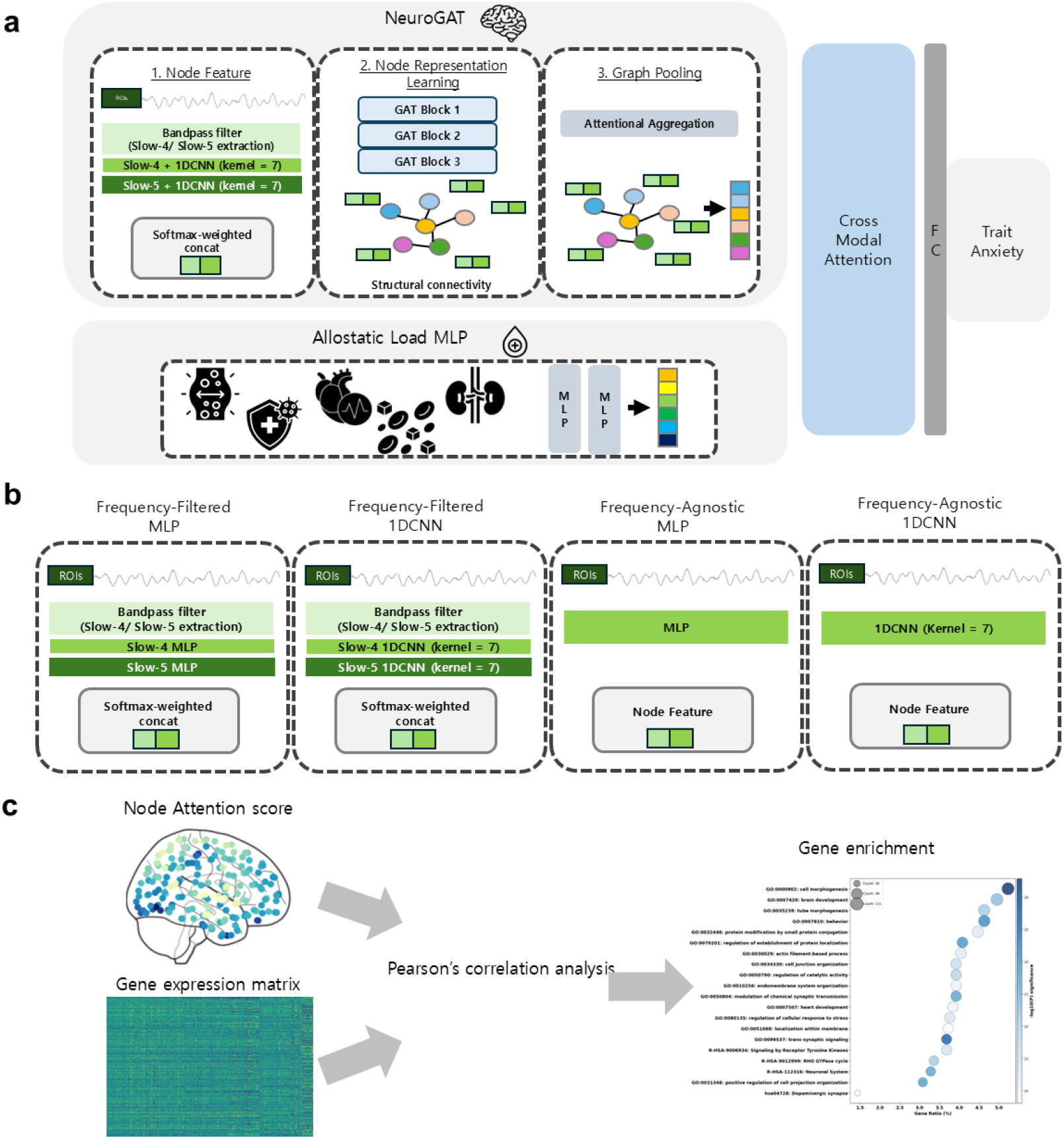
Overview of the analytical framework. **a. Model Architecture.** The framework integrates rs-fMRI–derived neural features and diffusion MRI–based structural connectivity via a graph attention network (NeuroGAT), combines them with physiological allostatic load markers representations through cross-modal attention and predicts trait anxiety. **b. Node Feature Extraction**. Four strategies were implemented to test the contribution of temporal dynamics and frequency information in fMRI signals (frequency-filtered MLP, frequency-filtered 1DCNN, frequency-agnostic MLP and frequency-agnostic 1DCNN). **c. Gene Enrichment Workflow**. Attention scores from NeuroGAT highlight predictive brain regions, which are spatially correlated with the Allen Human Brain Atlas transcriptome and analyzed for functional enrichment.

## 2 Results

Our framework predicts individual trait anxiety outcomes (State-Trait Anxiety Inventory (STAI) [37, 38]) by integrating brain and physiological data (**Figure 1a**). To systematically assess the contribution of temporal and spectral information from rs-fMRI signals, we compared four node-feature extraction strategies within the same architecture (**Figure 1b**): (1) Frequency-filtered MLP, which uses slow-4 and slow-5 band-filtered inputs but ignores temporal ordering; (2)Frequency-filtered 1DCNN, which applies temporal convolution to the same band-limited signals (3) Frequency-agnostic MLP, which receives unfiltered signals without explicit frequency emphasis or temporal modeling and (4) Frequency-agnostic 1DCNN, which applies temporal convolution directly to unfiltered inputs, capturing temporal patterns but not frequency-specific content.

### 2.1 Temporal Dynamics Contribute to Trait Anxiety Prediction

A one-way ANOVA revealed significant performance differences across node feature extraction strategies for all evaluation metrics (MSE: *p* = 0.0336; *r*^2^: *p* = 0.0401; *r*: *p* = 0.0132). Tukey’s post-hoc tests indicated that the frequency-filtered 1DCNN significantly outperformed both the frequency-filtered MLP (*p* = 0.0362) and the frequency-agnostic MLP (*p* = 0.0233), showing that incorporating temporal dynamics within slow oscillation bands (*slow-4*/*slow-5*) significantly improves trait-anxiety prediction compared with models that disregard temporal order (**Figure 2a**).

**Figure 2.**
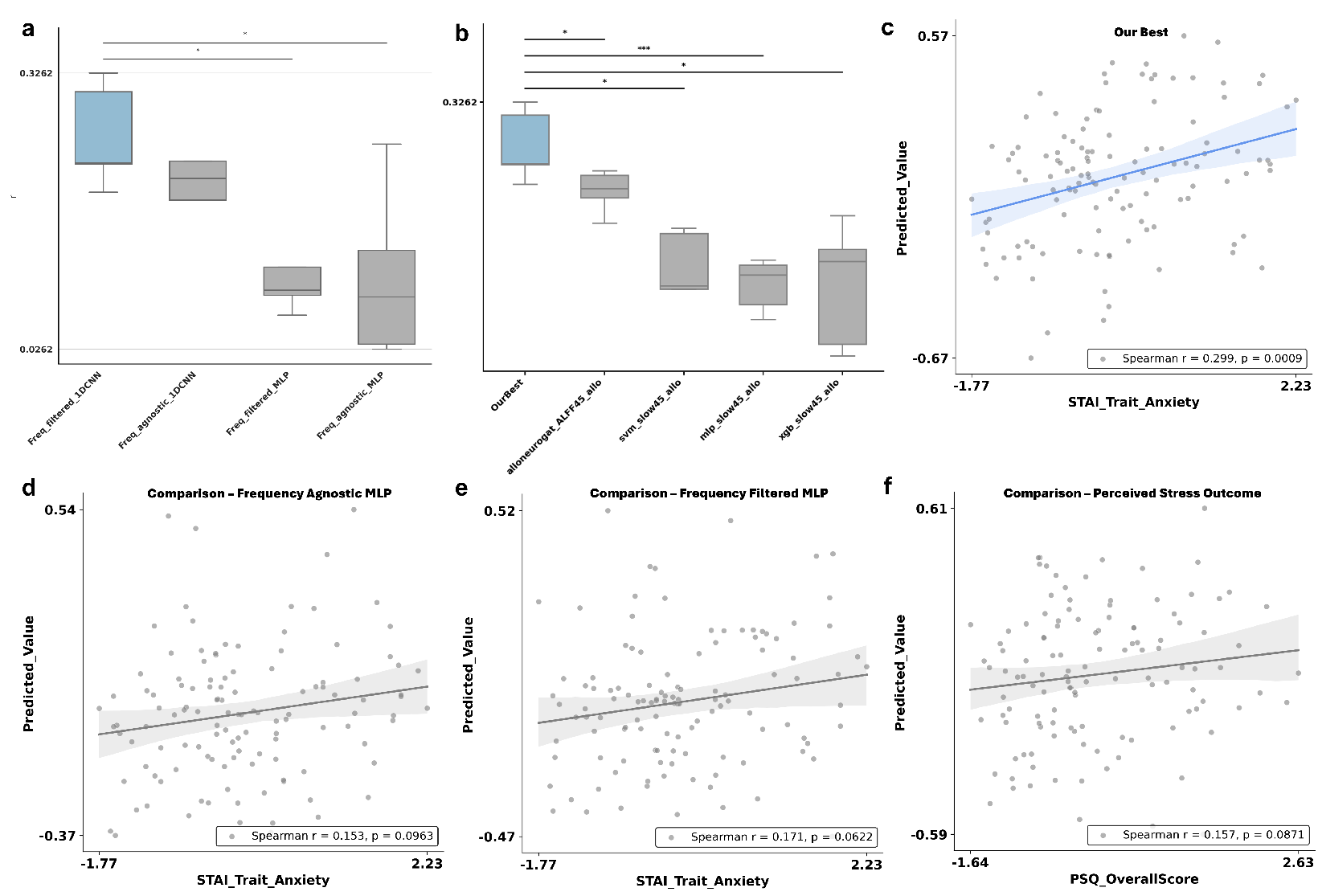
Incorportating Temporal Dynamics Improves Model Performance for Trait Anxiety Prediction. **a. Comparison of node feature extraction strategies.** One-way ANOVA followed by Tukey post-hoc tests compared four node feature extraction methods. The frequency-filtered 1DCNN model significantly outperformed frequency-agnostic MLP models in Pearson correlation (*r*), highlighting the importance of temporal dynamics. **b. Comparison with baseline models**. The proposed model achieved significantly higher correlations than conventional models (SVM, XGBoost and MLP) trained on ROI-wise Amplitude of Low-Frequency Fluctuations (ALFF) (slow-4 and slow-5 bands) and physiological markers, demonstrating superior predictive performance. **c. Ensemble prediction of trait anxiety (best model)**. Predicted values were averaged across five repeated runs for each participant and correlated with observed trait anxiety scores (Spearman *rho* = 0.299, *p* = 0.0009). No significant sex differences were observed in predicted values (two-sample *t*-test, *p* = 0.6165; see Supplementary Material). **d. Ensemble prediction using frequency-agnostic MLP features**. Same analysis as in panel c but using frequency-agnostic MLP node features. **e. Ensemble prediction using frequency-filtered MLP features**. Same analysis as in panel c but using frequency-filtered MLP node features. **f. Ensemble prediction for Perceived Stress using freqeuncy-filtered 1DCNN**. Same analysis as in panel c but using frequency-filtered 1DCNN node features for predicting Perceived Stress instead of Trait Anxiety.

We next compared the proposed frequency-filtered 1DCNN with models trained on conventional amplitude-based features. When node features were replaced by *slow-4* and *slow-5* ALFF values, performance significantly declined (paired *t*-test: MSE *p* = 0.024; *r*^2^ *p* = 0.024; *r p* = 0.045). Likewise, classical machine-learning models trained on ALFF and allostatic-load features (SVM, XGBoost, MLP) yielded negative *r*^2^ values (**Figure 2b**). These results indicate that the performance gap is driven by the presence of temporally resolved neural features rather than the choice of learning algorithm: when inputs lack temporal dynamics, both shallow and deep models perform poorly.

To comprehensively assess model specificity and multimodal contributions, we performed additional comparison experiments. Perceived stress was chosen as a reference phenotype for testing specificity because it indexes transient stress responses to environmental demands, whereas trait anxiety reflects a stable, trait-like vulnerability [39]. If the model captures general stress reactivity rather than enduring predispositions, it should predict perceived stress equally well. However, predictive performance was significantly lower for perceived stress (Perceived Stress Questionnaire [40]; Trier Inventory for Chronic Stress [41]; PSQ: MSE *p* = 0.021, *r*^2^ *p* = 0.021, *r p* = 0.007; TICS: MSE *p* = 0.029, *r*^2^ *p* = 0.029, *r p* = 0.014), confirming that the learned representations capture biological signatures specific to trait anxiety rather than transient stress states.

Comparison against unimodal baselines revealed a significant difference in pearson correlation performance metric (**Table1̃**; ANOVA *p* = 0.0002). Post-hoc Tukey tests showed that the multimodal model significantly outperformed the brain-representation–only variant (*p* = 0.0002), while its difference from the allostatic-load–only variant was not significant. Nonetheless, the multimodal model showed slightly lower variance across repetitions (Levene’s test *p* = 0.067) than the physiology-only variant, indicating more stable generalization. These findings indicate that while allostatic-load markers alone explain part of the variance in trait anxiety, integrating neural features enhances robustness and complementary representation. Detailed ablation analyses varying attention configurations and connectivity inputs are presented in the Supplementary Materials.

**Table 1:** Model Comparison of comparison models.

| Model | MSE (↓) | $r^2$ (↑) | Pearson $r$ (↑) |
| --- | --- | --- | --- |
| ALFF (XGBOOST) | $1.208 \pm 0.099$ | $-0.217 \pm 0.124$ | $0.037 \pm 0.087$ |
| ALFF (MLP) | $1.201 \pm 0.033$ | $-0.208 \pm 0.054$ | $0.037 \pm 0.037$ |
| ALFF (SVM) | $1.090 \pm 0.057$ | $-0.080 \pm 0.066$ | $0.041 \pm 0.084$ |
| AlloNeuroGAT-Freq_agnostic_MLP | $1.005 \pm 0.051$ | $-0.030 \pm 0.052$ | $0.105 \pm 0.082$ |
| AlloNeuroGAT-Freq_filtered_MLP | $1.008 \pm 0.060$ | $-0.034 \pm 0.061$ | $0.115 \pm 0.056$ |
| NeuroGAT Only - Freq_filtered_1DCNN | $0.986 \pm 0.022$ | $-0.012 \pm 0.023$ | $-0.068 \pm 0.081$ |
| AlloNeuroGAT-ALFF | $0.964 \pm 0.011$ | $0.012 \pm 0.011$ | $0.186 \pm 0.030$ |
| AlloOnly-AL_MLP | $0.948 \pm 0.051$ | $0.028 \pm 0.052$ | $0.187 \pm 0.095$ |
| AlloNeuroGAT-Freq_agnostic_1DCNN | $0.940 \pm 0.035$ | $0.036 \pm 0.036$ | $0.206 \pm 0.072$ |
| **AlloNeuroGAT-Freq_filtered_1DCNN** | **$0.924 \pm 0.028$** | **$0.052 \pm 0.028$** | **$0.257 \pm 0.050$** |

Under ensemble averaging across five trained folds, only the frequency-filtered 1DCNN showed a significant correlation with trait anxiety (*r*_*s*_ = 0.299, *p* = 0.0009; **Figure 2c**), whereas MLP ensembles did not (**Figures 2d–e**). Ensemble predictions for perceived stress were non-significant (*r*_*s*_ = 0.157, *p* = 0.0871; **Figure 2f**), supporting trait-specific rather than state-dependent modeling effects.

### 2.2 Limbic and Visual Networks Emerge as Core Predictive Hubs

We next analyzed which brain connections contributed most strongly to the individual-level prediction of trait anxiety. In the graph attention framework, each edge is assigned an attention value (**Figure 3a**; Methods) and node-level importance was derived by averaging attention across all incident edges (**Figure 3b**).

**Figure 3.**
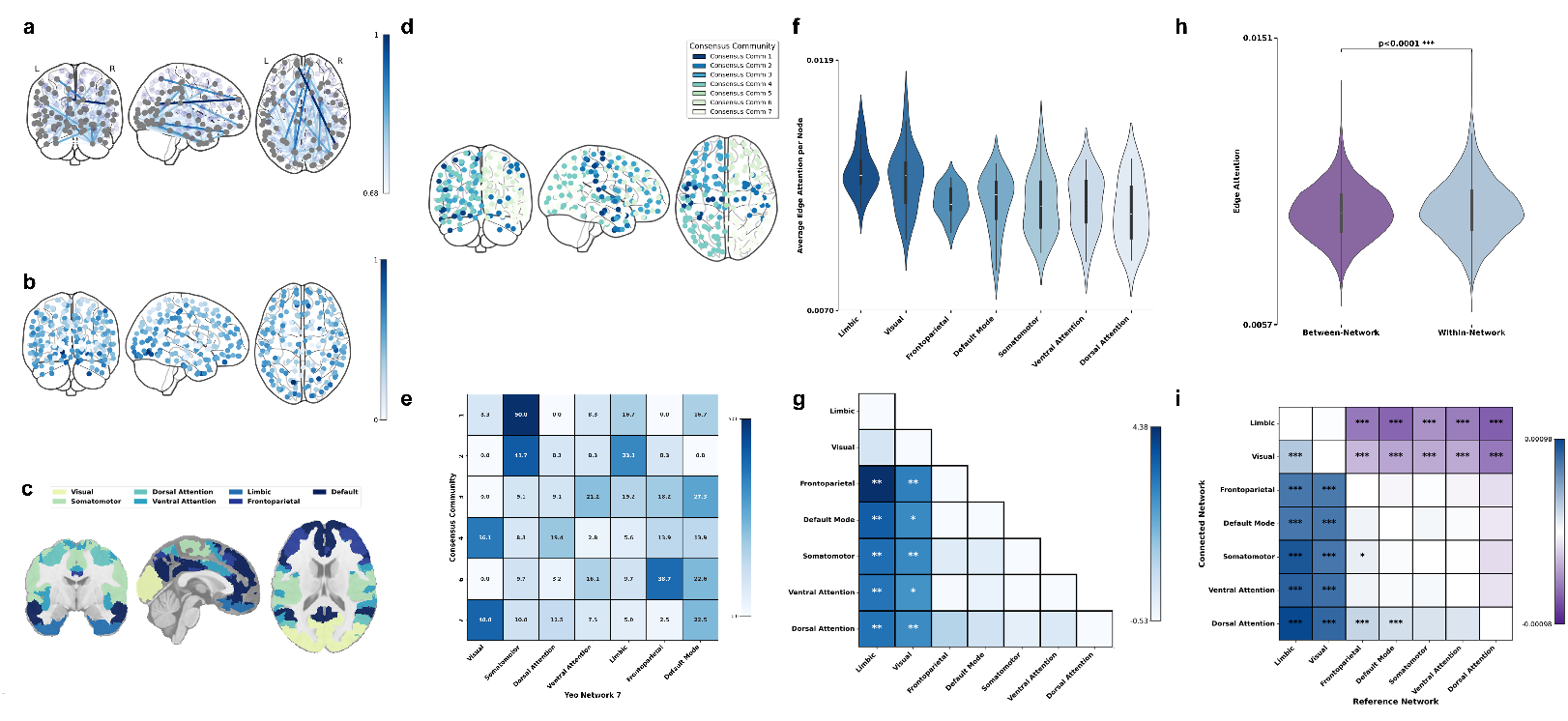
Predictive Attention value for Trait Anxiety Converges in Limbic and Visual Networks. **a. Visualization of the most highly attended edge connections.** Top 1% of edges ranked by mean attention weights, scaled by value, highlighting the most predictive structural connections. **b. Node importance computed as the average attention of incident edges. c. Visualization of Yeo 7 Functional Network d. Consensus communities:** Seven communities were identified using the Leiden algorithm (150 runs). This matrix was clustered using agglomerative clustering across candidate community numbers (2–10) and the solution with the highest silhouette score (0.9182) was chosen as the final consensus partition. **e. Overlap with Yeo networks:** Heatmap showing percentage overlap between consensus communities and Yeo 7 functional networks (observed Mutual Information = 0.3122, permutation *p <* 0.00001); darker shades indicate higher overlap (Community 5, consisting of a single node, was excluded). **f. Distribution of node attention across Yeo 7 networks**. A Kruskal–Wallis test (*H* = 31.03, *p <* 0.0001) revealed significant differences. **g. Post-hoc analysis of attention scores across Yeo 7 functional network**. Post-hoc Dunn’s tests (FDR-BH corrected, *α* = 0.05) showed the Limbic and Visual networks carried significantly higher attention than most other networks (e.g., Limbic vs. Default Mode *p*_*corrected*_ = 0.0026; Visual vs. Default Mode *p*_*corrected*_ = 0.0109). No difference was found between Limbic and Visual (*p*_*corrected*_ = 0.5753). **h. Within-vs. between-network edge attention**. Within-network edges (*N* = 1950) showed significantly higher attention than between-network edges (*N* = 11, 044, Mann–Whitney U, *p <* 0.0001). **i. Within- and between-network attention weights in Yeo network level**. Heatmap of within–between network differences in mean attention weights. Blue = stronger within-network attention; purple = stronger between-network attention. Significance assessed by Mann–Whitney U with FDR-BH correction (\**p*_*corrected*_ *<* 0.05, \*\**p*_*corrected*_ *<* 0.01, \*\*\**p*_*corrected*_ *<* 0.001).

To test whether attention-derived topology reflects known large-scale organization, we performed consensus community detection on node-level attention maps using 150 runs of the Leiden algorithm. The final partition was selected by maximal silhouette score (0.9182). Mutual information analysis revealed significant correspondence between attention-derived communities and Yeo 7 functional networks (**Figure 3c**) [42] (observed MI = 0.3122, permutation *p <* 0.00001; **Figure 3d**). The resulting communities aligned strongly with canonical systems, including somatomotor (communities 1 and 2: 50% and 41.7%), visual (communities 4 and 7: 36.1% and 40.0%) and frontoparietal (community 6: 38.7%), demonstrating that the model’s learned representations align with macroscale functional organization rather than arbitrary structure (**Figure 3e**).

To contextualize node-level attention within canonical systems, nodes were grouped by Yeo 7 networks. Attention distributions differed significantly among networks (Kruskal–Wallis, *p <* 0.0001; **Figure 3f**). FDR-corrected post-hoc comparisons indicated significantly higher attention within the limbic (*p*_*corrected*_ *<* 0.01) and visual (*p*_*corrected*_ *<* 0.05) networks relative to other systems (**Figure 3g**), indicating that predictive information for trait anxiety is concentrated in these two networks rather than being broadly distributed across the brain.

We next examined how predictive edges were organized across and within functional systems. Intra-network connections carried significantly higher attention value than inter-network connections overall (**Figure 3h**; Mann–Whitney U, *p <* 0.0001), suggesting that locally cohesive circuits play a key role in trait anxiety prediction. This pattern was primarily driven by the limbic and visual networks, which emerged as dominant sources of intra-network integration. At the network level, the limbic system showed stronger intra-network attention value than any of its inter-network connections (*p*_*corrected*_ *<* 0.001 for all comparisons) and the visual network exhibited similarly showed higher within-network attention except for its connections with the limbic system (*p*_*corrected*_ *<* 0.001). Conversely, other networks showed higher attention for edges linking to the limbic and visual systems than for their own intra-network connections (**Figure 3i**). Together, these findings indicate that the limbic and visual systems act as central predictive hubs, contributing strongly to both within-network integration and cross-network connectivity with other large-scale systems.

### 2.3 Dynamic Functional Connectivity and Association with Trait Anxiety

To further probe the functional relevance of temporal dynamics, we estimated dynamic functional connectivity (dFC) states from sliding-window correlations of resting-state BOLD signals. Analyses were conducted separately for the *slow-4* (0.027–0.073 Hz) and *slow-5* (0.01–0.027 Hz) bands. Elbow analysis indicated that four recurrent connectivity states (*k* = 4) best captured temporal variance in both frequency bands (**Figure 4a,f**; k=3 and 5 in Supplementary Materials). Each participant’s windows were reassigned to these state centroids and state-mean FC matrices were computed by averaging all ROI–ROI correlations belonging to each state. The upper panels of **Figure 4a,b** show these ROI-level mean FC maps, while the lower panels depict network-level coupling averaged within and between Yeo-7 functional systems.

**Figure 4.**
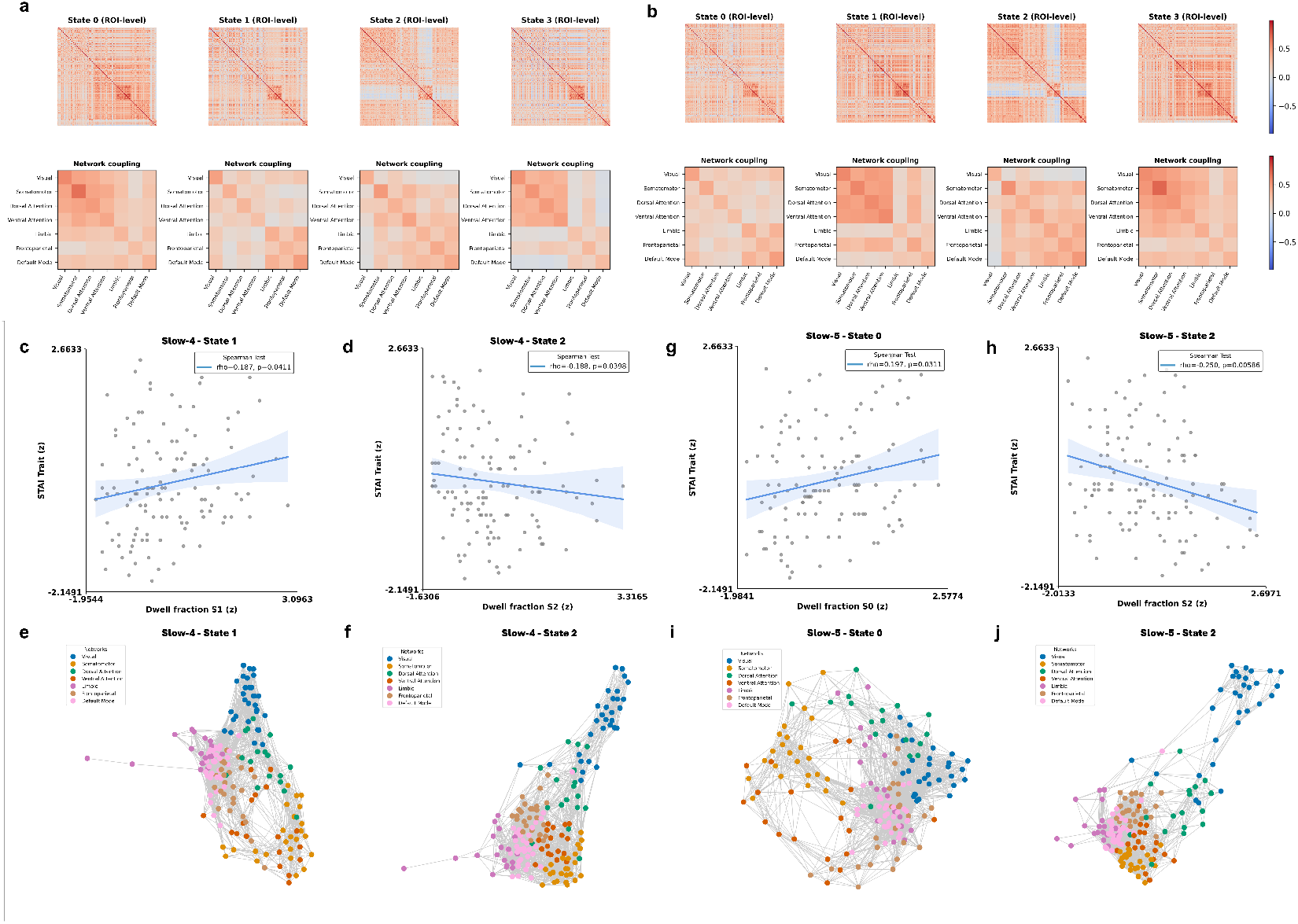
Dynamic Functional Connectivity (dFC) States and State Dwell-Fraction Association with Trait Anxiety. **a. ROI- and network-level representations of the four dFC states identified in the *slow-4* band.** Upper panels show ROI-level mean FC matrices; lower panels show network-level coupling averaged within and between Yeo-7 networks. **b. ROI- and network-level representations of the four dFC states identified in the *slow-5* band**. Overall state topologies were highly similar to those in *slow-4*, as confirmed by cross-band similarity analysis using Euclidean distance (1*/*(1 + distance)), which revealed one-to-one correspondences between matched state pairs (Slow4_State_0–Slow5_State_3: 0.0245; Slow4_State_1–Slow5_State_0: 0.0238; Slow4_State_3– Slow5_State_1: 0.0219; Slow4_State_2–Slow5_State_2: 0.0196). **c. Positive association between the dwell fraction of State 1 (slow-4) and trait anxiety**. Longer dwell time compared to total time points in State 1 was correlated with trait anxiety (*rho* = 0.187, *p* = 0.0411). **d. Negative association between the dwell fraction of State 2 (slow-4) and trait anxiety**. Shorter dwell time compared to total time points in State 2 was correlated with trait anxiety (*rho* = *−* 0.188, *p* = 0.0398). **e–f. Graph-theoretical representations of State 1 (e) and State 2 (f) in the *slow-4* band, thresholded at the top 20% of positive correlations**. Nodes represent ROIs colored by Yeo-7 network membership; edges denote strong functional links. **g. Positive association between the dwell fraction of State 0 (slow-5) and trait anxiety**. Longer dwell time compared to total time points in State 0 was correlated with trait anxiety (*rho* = 0.197, *p* = 0.0311). **h. Negative association between the dwell fraction of State 2 (slow-5) and trait anxiety**. Shorter dwell time compared to total time points in State 2 was correlated with trait anxiety (*rho* = *−* 0.250, *p* = 0.0059). **i–j. Graph-theoretical representations of State 0 (i) and State 2 (j) in the *slow-5* band, thresholded at the top 20% of positive correlations**.

Despite being derived from distinct frequency bands, state topologies were highly consistent across *slow-4* and *slow-5*. Euclidean-distance–based similarity analysis (*S* = 1*/*(1 + *∥x −y ∥*_2_)) revealed clear one-to-one correspondences between matched states (Slow4_State_0–Slow5_State_3 = 0.0245; Slow4_State_1–Slow5_State_0 = 0.0238; Slow4_State_3–Slow5_State_1 = 0.0219; Slow4_State_2–Slow5_State_2 = 0.0196). Mantel permutation testing (5,000 iterations) confirmed significant structural correspondence (*p* = 0.0002 for all pairs), indicating frequency-invariant large-scale network configurations (**Figure 4a,b**).

We next examined behavioral relevance by testing the association between each state’s dwell fraction and trait anxiety. In the *slow-4* band, longer dwell time compared to total time points in State 1 and shorter dwell time in State 2 were associated with higher trait anxiety (Spearman *rho* = 0.187, *p* = 0.0411; *rho* = *−* 0.188, *p* = 0.0398; **Figure 4c,d**). To interpret these associations, we inspected the corresponding network topologies utilizing (see Methods). Graph-theoretical visualizations thresholded at the top 20% of positive correlations revealed that State 1 was characterized by strong coupling among limbic, default-mode and frontoparietal networks (**Figure 4e**), whereas State 2 exhibited lower global integration with the visual network relatively segregated from other systems (**Figure 4f**).

Notably, similar patterns emerged in the *slow-5* band. Its State 0 and State 2, structurally corresponding to *slow-4* State 1 and State 2, respectively, showed identical behavioral associations, where greater dwell time compared to total time points in State 0 and lower dwell time in State 2 predicted higher trait anxiety (*rho* = 0.197, *p* = 0.0311; *rho* = *−*0.250, *p* = 0.0059; **Figure 4g,h**). Network topology analyses confirmed parallel configurations: State 0 displayed pronounced limbic–default-mode–frontoparietal coupling, whereas State 2 showed relative decoupling of the visual network from the rest of the brain (**Figure 4i,j**).

Together, these results indicate that individuals with higher trait anxiety tend to spend more time in states marked by strong limbic–default-mode-frontoparietal integration and less time in states where the visual network is functionally detached.

### 2.4 Physiological Biomarkers Capture Variability in Trait Anxiety

We also assessed the relative importance of physiological markers. Permutation-based feature importance scores (see Methods), estimated by shuffling each marker and quantifying the resulting drop in model performance, revealed that metabolic and immune markers, including creatinine (CRE), Body Mass Index (BMI), glucose (GLU), total cholesterol-to-high-density lipoprotein ratio (CHOL_HDLC_ratio) and C-reactive protein (CRP), contributed strongly to prediction (**Figure 5a**), whereas cardiovascular indices were consistently weaker predictors. Spearman correlation analyses (**Figure 5b**) further supported these findings, revealing significant associations with trait anxiety after FDR-BH correction: negative with CRE (*rho* = *−*0.298, *p*_corrected_ = 0.0095) and BMI (*rho* = *−*0.264, *p*_corrected_ = 0.018) and positive with GLU (*rho* = 0.241, *p*_corrected_ = 0.027).

**Figure 5.**
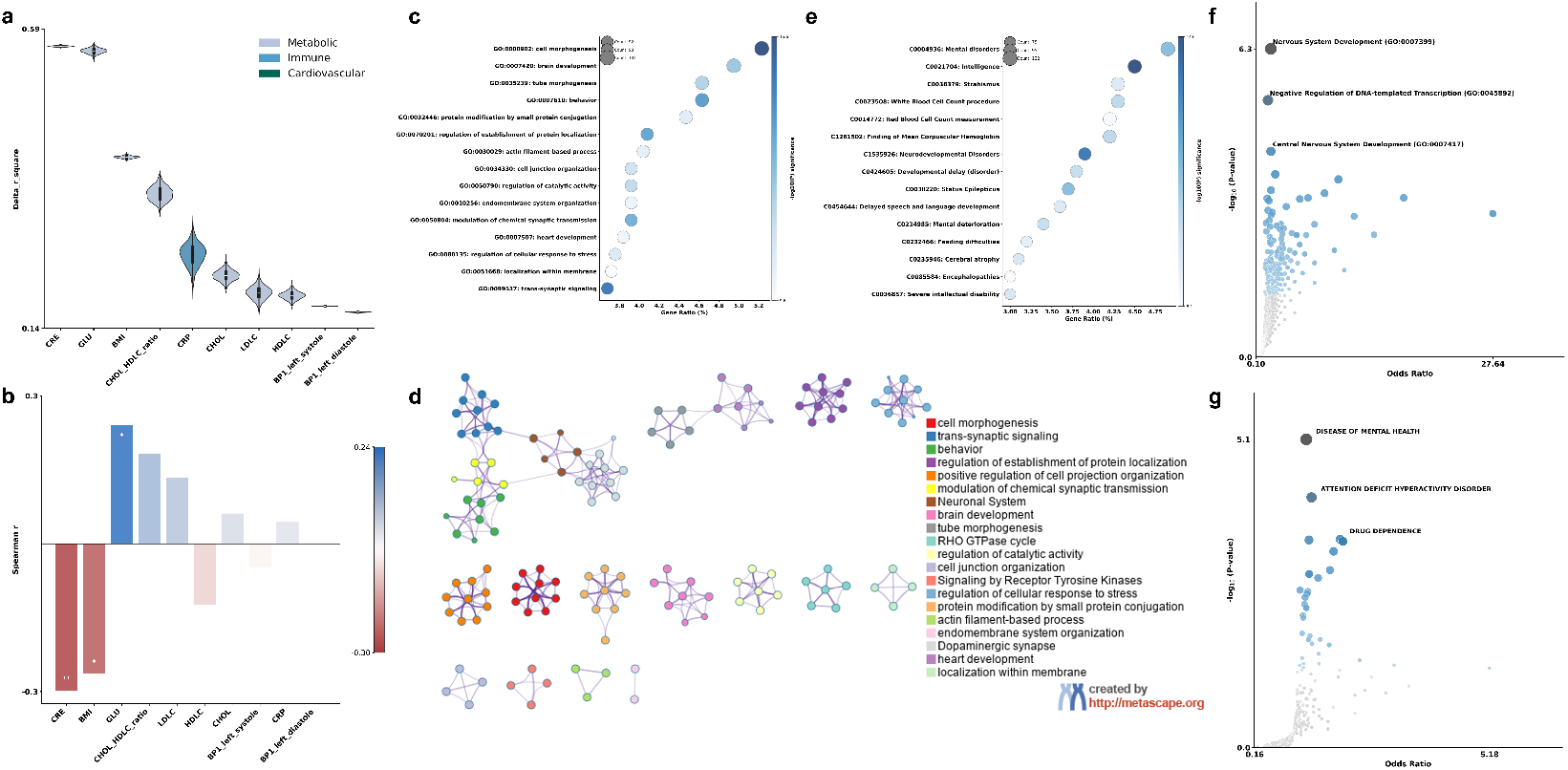
Importance of Physiological Markers and Transcriptomic Signatures Derived from Attention-Based Node Importance. **a. Physiological marker importance.** Permutation-based feature importance revealed strong contributions of metabolic and immune markers (CRE, GLU, BMI, CHOL/HDLC ratio, CRP). **b. Correlation with anxiety**. Spearman correlations showed significant associations of CRE (negative), BMI (negative) and GLU (positive) with trait anxiety after FDR-BH correction. **c. Functional enrichment of attention-correlated genes**. Bubble plot of the top 15 enriched Gene Ontology (GO) and pathway terms utilizing Metascape. The x-axis shows the proportion of genes within each term (Gene Ratio), bubble size indicates the number of genes and bubble color encodes significance (*−* log_10_ p-value). Enriched terms highlight processes related to neurodevelopment, synaptic signaling and behavioral regulation. **d. Enrichment network of functionally related terms**. Each node represents an enriched GO term, colored by functional category. Edges connect terms with similarity scores *>*0.3, forming clusters of biologically related processes. A major interconnected module centers on neuronal signaling and synaptic processes, with additional modules capturing morphogenesis and cellular regulation. **e. Disease enrichment of attention-correlated genes**. Bubble plot showing the top 15 enriched DisGeNET terms. As in panel c, the x-axis represents gene ratio, bubble size reflects gene count and color intensity corresponds to significance. Significant associations were observed for mental and neurodevelopmental conditions (e.g., mental disorders, developmental delay, neurodevelopmental disorders), underscoring the clinical relevance of trait anxiety. **f. Volcano Plot of terms from the GO Biological Process 2025**. Each point represents a single gene ontology term from the GO Biological Process 2025 library on Enrichr, plotted by the corresponding odds ratio (x-position) and *−* log_10_(*p*-value) (y-position). The larger and darker-colored the point, the more significantly enriched the input gene set is for the term. The top three most significantly enriched terms, based on the *−* log_10_(*p*-value), are labeled in the figure. **g. Volcano Plot of terms from the Jensen DISEASES Experimental 2021**. Enrichment analysis using the same input gene set as in panel d, but querying the Jensen DISEASES Experimental 2021 library on Enrichr. Each point represents a single term, plotted by its odds ratio (x-axis) and *−* log_10_(*p*-value) (y-axis). The top three most significantly enriched terms are labeled.

### 2.5 Transcriptomic Pathways Ground the Network Signature

To connect the model-derived network signature with underlying molecular architecture, we correlated regional node attention scores with spatial gene-expression profiles from the Allen Human Brain Atlas. Genes showing significant associations after Bonferroni correction(*p*_*corrected*_ *<* 0.05) were compiled into gene set enrichment analysis (GSEA) using Metascape [43]. Non-redundant GO modules converged on three dominant biological themes: (i) developmental and morphogenetic programs, (ii) synaptic signaling and neuronal communication and (iii) cellular regulation and stress-response pathways (**Figure 5c**). Representative terms such as *cell morphogenesis* (*q <* 10^*−*12.5^), *trans-synaptic signaling* (*q <* 10^*−*11.5^) and *brain development* (*q <* 10^*−*9.5^), underscoring strong enrichment for developmental processes that shape brain structure. The second major axis encompassed synaptic and neuronal communication. Gene ontology terms linked to *trans-synaptic signaling* included neurotransmitter secretion, synaptic vesicle exocytosis and regulated exocytosis. The *modulation of chemical synaptic transmission* cluster captured regulation of trans-synaptic signaling, synaptic plasticity and long-term potentiation. The *dopaminergic synapse* was embedded in a broader neurotransmission network that included cholinergic, glutamatergic, GABAergic and serotonergic signaling, as well as endocannabinoid and circadian pathways. The *neuronal system* module encompassed NMDA receptor–mediated signaling. Importantly, these neurotransmission modules were not isolated but showed direct connections with the *behavior* cluster related to cognition, learning, memory and associative or visual learning. (**Figure 5d**).

The third axis consisted of cellular regulatory and stress-related processes. Gene ontology such as *regulation of catalytic activity* (*q <* 10^*−*8.7^), *regulation of establishment of protein localization* (*q <* 10^*−*10.5^), *protein modification by small protein conjugation* (*q <* 10^*−*7.7^) and *regulation of cellular response to stress*(*q <* 10^*−*7.9^) were included.

Disease enrichment analysis using DisGeNET further underscored the clinical relevance of these genes (**Figure 5e**). Strong enrichment was observed for mental and neurodevelopmental disorder categories, with significant associations also found for phenotypes such as delayed speech and language development.

To confirm the robustness of the molecular enrichment, we conducted an independent Enrichr analysis across multiple databases[44]. The results converged on neurodevelopmental and synaptic signaling processes, with significant terms including nervous system development (*GO BP 2025, q <* 10^*−*2.7^; **Figure 5f**), dopaminergic synapse (*KEGG 2021, q <* 10^*−*2.8^) and BDNF signaling pathway (*BioPlanet 2019, q <* 10^*−*1.9^). Cellular-level enrichment for axonal growth cone (*GO CC 2025, q <* 10^*−*1.8^) and disease-level enrichment for mental health disorders (*Jensen Diseases Experimental 2025, q <* 10^*−*2.5^; **Figure 5g**) further support the neurodevelopmental and neurotransmission-related nature of the identified gene set.

## 3 Discussion

This study examined multiscale biological signatures of trait anxiety by integrating temporal brain dynamics and systemic allostatic markers within a multimodal deep-learning framework. Temporal modeling of slow-frequency rs-fMRI signals significantly improved prediction over static or temporally unordered models, underscoring the role of dynamic neural information in stable anxiety vulnerability. The multimodal model also significantly outperformed the brain-only variant and showed marginally greater stability than the allostatic-only model. Attention-based interpretation highlighted the limbic and visual networks as key predictive systems. Individuals with higher trait anxiety spent more time in brain states marked by limbic–default-mode–frontoparietal hyperintegration and less time in visually segregated states. Metabolic and immune markers, including creatinine, glucose and BMI, further contributed to prediction, linking neural dynamics to peripheral physiology. Transcriptomic enrichment revealed that anxiety-relevant network hubs aligned with genes involved in neurodevelopment, synaptic signaling—particularly dopaminergic pathways—and stress response. Collectively, these findings characterize trait anxiety as a brain–body phenomenon rooted in dysregulated limbic–visual circuit dynamics.

Previous studies have reported that the amplitude of slow-4 and slow-5 oscillations (ALFF) is associated with anxiety [29, 45, 46]. However, we quantitatively demonstrate temporal dynamics within these low-frequency bands play a decisive role in predicting individual differences in trait anxiety. Models capturing temporal dynamics within slow oscillations markedly outperformed those based on static amplitude or temporally unordered inputs, indicating that anxiety vulnerability is embedded not in momentary hyper- or hypoactivation but in the temporal organization of neural activity over time. This shift in focus aligns with recent advances highlighting temporal properties of resting-state fMRI in explaining attention, perceptual and cognitive variability [47–51].

Nevertheless, temporal neural dynamics alone were insufficient for stable prediction. Integrating them with systemic physiological markers yielded higher validity and marginally better reproducibility, suggesting that anxiety vulnerability emerges from the interaction between neural temporal variability and allostatic systems. This interpretation is consistent with evidence that physiological factors can substantially influence functional connectivity measures [52]. Moreover, experimental work has demonstrated this relationship directly: inducing peripheral inflammation altered both the temporal variability of resting-state BOLD signals and functional connectivity within emotion-regulation regions [53], indicating that physiological perturbations can modulate neural dynamics in a measurable way. Together, these findings highlight that the brain and body function as an integrated system and that such coupling offers a more comprehensive account of anxiety predisposition. Finally, our model predicted trait anxiety but not perceived stress, reinforcing that these neural–physiological signatures reflect enduring vulnerability traits rather than general stress reactivity.

The analyses utilizing attention values from graph attention neural network identified the limbic and visual networks as key predictive hubs for trait anxiety. These two systems occupy opposite poles of the cortical hierarchy [54]: the limbic network, deeply embedded and affectively oriented and the visual network, positioned at the perceptual interface. Despite their hierarchical distance, both showed high predictive importance. Furthermore, our model revealed how these systems function as hubs: they were dominated by strong importance within intra-network, while other higher-order networks derived their predictive importance primarily from their inter-network connections *to* these two core networks. The limbic network is known to receive dense subcortical projections from regions such as the amygdala and hippocampus [55] and plays a central role in emotional salience and valuation [56–60]. Its cortical extensions into orbitofrontal and anterior temporal areas [61] support affective meaning and social-context processing. Visual network involvement in anxiety has not been a major focus in prior research, but several studies have reported hyperactivation to threat-related cues within visual regions [62, 63]. Taken together, these attention-based findings point to the limbic and visual circuits as key functional networks for understanding trait anxiety. Their opposing positions within the cortical hierarchy suggest that vulnerability may arise from altered coordination between affective and perceptual systems.

To assess whether model-derived temporal information reflects functionally relevant dynamics, we estimated recurrent dynamic functional connectivity states from resting-state fMRI. Resting-state BOLD activity is known to exhibit quasi-periodic fluctuations [64] and we hypothesized that these temporal variations may capture intrinsic features of anxiety-related network organization. Individuals with higher trait anxiety spent more time in states characterized by strong coupling among the limbic, default mode and frontoparietal networks (State 0/1 ; slow-4 and slow-5) and less time in states where the visual network was functionally segregated from other systems (State 2; both frequency bands). These results suggest a tendency for anxious individuals to remain “locked” in internally focused, affectively integrated configurations, while failing to maintain transiently segregated visual states that may support perceptual disengagement and sensory rest.

This pattern aligns closely with the Attentional Control Theory of anxiety [65], which attributes anxiety-related deficits to (1) enhanced capture by perceptual distractors and (2) impaired inhibition in shifting attention once captured. The lower dwell time in visually segregated states observed here may indicate insufficient down-regulation of sensory input, consistent with behavioral findings of heightened perceptual sensitivity under high perceptual load in anxiety [66, 67]. Conversely, the prolonged coupling among limbic, DMN and FPN networks reflects difficulty in control switching. Typically, the FPN supports goal-directed external attention, while the DMN mediates self-referential processing and their activity is anticorrelated under healthy conditions [68, 69]. Prior studies have shown that this anticorrelation breaks down in anxiety and depression, yielding increased positive coupling [70–72] and that individuals with generalized anxiety disorder exhibit difficulty transitioning away from such coactivation states to other state [73]. Our findings extend these observations by demonstrating this neural stickiness dimensionally across healthy individuals, linking higher trait anxiety to persistent internal coupling and reduced sensory segregation. Together, these findings provide neural-dynamic evidence supporting the core assumptions of Attentional Control Theory.

Our study further demonstrated that metabolic and immune markers constitute major physiological axes explaining individual variability in trait anxiety. Permutation-based importance and correlation analyses revealed that metabolic indicators, including creatinine (CRE), body mass index (BMI) and glucose (GLU), were significantly associated with anxiety levels, highlighting the metabolic–immune system as a core contributor to anxiety vulnerability.

The negative association between CRE and anxiety is particularly noteworthy. Creatinine is known to cross the blood–brain barrier and be taken up by neurons and oligodendrocytes in the hippocampus and cortex, where it supports neuronal energy metabolism [74, 75]. This mechanism aligns with reports of lower brain creatine concentrations in patients with anxiety and post-traumatic stress disorder [76–78]. In contrast, elevated glucose levels have been consistently linked to metabolic dysregulation and heightened stress reactivity in anxious populations [79, 80] and our findings suggest that such metabolic vulnerability is closely tied to the neural substrates of anxiety. The relationship between BMI and anxiety has been inconsistent across studies, with some reporting U-shaped associations [81, 82]. The negative correlation observed in our sample is likely attributable to its distribution toward the underweight–normal range (see Supplementary Materials), suggesting that limited energy availability or altered physiological stress reactivity may influence this direction of association. Although C-reactive protein (CRP) did not show direct correlations with anxiety, it ranked highly in permutation-based importance, supporting the role of chronic low-grade inflammation as a potential physiological substrate of anxiety. By contrast, cardiovascular indices showed consistently low contributions, suggesting that trait anxiety may be more closely related to long-term dysregulation of metabolic and immune axes [83] than to acute arousal-related responses. Taken together, these findings indicate that anxiety vulnerability may emerge from imbalances in metabolic and immune regulation alongside temporal neural dynamics. Rather than reflecting hyperactivity within a single neural system, anxiety appears to arise from an interactive framework linking energy metabolism, immune signaling and neural activity.

Building on this foundation, we mapped high-attention regions to AHBA transcriptomics to derive molecular context of trait anxiety. Enrichment converged on three axes (1) neurodevelopmental and morphogenetic programs, (2) synaptic signaling and neuronal communication and (3) cellular regulation and stress-response pathways. The synaptic axis involved dopaminergic, glutamatergic, cholinergic, and GABAergic signaling, linking molecular communication to cognition and learning, while developmental and regulatory programs supported circuit formation and stress adaptation. These findings are consistent with large-scale transcriptomic work showing shared dysregulation of neuronal and synaptic modules across multiple psychiatric disorders [84]. Interestingly, disease enrichment analyses (DisGeNET, Jensen Diseases Experimental 2025) and biological process annotations further highlighted mental and neurodevelopmental disorder categories, suggesting that the identified genes overlap with transdiagnostic neurodevelopmental pathways. Together, these convergent pathways align with the view that high trait anxiety reflects a core vulnerability with neurodevelopmental origins [85]. To confirm the robustness of the molecular enrichment, we conducted an independent GSEA using the Enrichr platform across multiple databases. This validation reinforced our primary findings, revealing consistent enrichment across dopaminergic and BDNF signaling, axonal growth cone components, and mental-health-related gene sets (*q <* 0.02). The consistent results across two distinct GSEA tools (Metascape and Enrichr) and various database libraries confirm that the transcriptomic signature is biologically grounded, linking synaptic signaling and developmental regulation to anxiety vulnerability.

The biological grounding of our model is noteworthy, as its core representational unit is derived from BOLD time-series data. Prior multimodal neuroimaging research integrating proton magnetic resonance spectroscopy with fMRI has shown that functional BOLD dynamics are sensitive to underlying neurochemical balance [86]. Within this context, the enrichment of our high-attention regions for dopaminergic synaptic genes is notable. Cortical D1 receptor mapping has revealed the highest receptor density in association networks such as the limbic and frontoparietal systems and the lowest in primary sensory areas like the visual cortex [87]. Together, these findings suggest that the state-specific coupling patterns observed in trait anxiety, particularly stronger limbic–default-mode–frontoparietal integration and reduced visual connectivity, may arise from neurotransmitter modulation or receptor distribution gradients that shape large-scale dynamic connectivity.

Despite these strengths, this study has several limitations. First, the predictive performance for trait anxiety did not reach a level sufficient for clinical application, which may reflect the fact that the cohort is composed primarily of healthy participants. Second, the AHBA transcriptomic data were derived from a limited number of postmortem samples (six individuals, predominantly left hemisphere, bulk-RNA-seq dataset), distinct from the imaging cohort, restricting interpretation at the level of individual variability and may limit generalization to the younger LEMON sample. Donor averaging may further obscure inter-individual variability relevant to anxiety. Third, physiological markers were measured at a single time point and thus may not fully capture day-to-day fluctuations or lifestyle influences. Finally, external validation was not feasible, as, to our knowledge, no publicly available dataset currently integrates resting-state fMRI, diffusion MRI, comprehensive blood biomarkers and trait anxiety measures simultaneously. Future research should (a) test the generalizability of these findings in longitudinal and clinical cohorts to evaluate their translational potential; (b) address individual variability (e.g., “brain fingerprint” [88–101]) in the context of trait anxiety.

In conclusion, the present study provides one of the few robustly designed deep learning frameworks that jointly integrate brain-body associations to predict anxiety vulnerability. The model elucidated which neural and physiological features hold predictive value, highlighting the complementary roles of temporal dynamics and metabolic–immune markers. Importantly, it identified the limbic and visual networks, often underemphasized in prior anxiety research, as central predictive hubs and empirically demonstrated their dynamic alterations as novel systems-level markers of vulnerability. Finally, by linking these network signatures to transcriptomic pathways related to neurodevelopment, synaptic signaling and stress response, this study advances a biologically grounded, multi-scale account of trait anxiety. Together, these results not only refine the mechanistic understanding of anxiety but also establish a foundation for future longitudinal and clinically oriented applications.

## Data Availability

All data produced are available online at https://fcon_1000.projects.nitrc.org/indi/retro/MPI_LEMON.html

## 6 Methods

### 6.1 Participants

Our study utilized data from the Max Planck Institute’s LEMON dataset [102], a comprehensive collection that includes resting-state functional magnetic resonance imaging (rsfMRI) time-series, diffusion MRI, blood biomarker information and behavioral data from healthy participants. Detailed recruitment procedures and exclusion criteria for the original dataset, including those for neurological/psychiatric conditions, medication use and substance abuse, are fully documented in its primary data release publication [102].

From this dataset, we included 120 young adults (84 males, age range 20-30 years) who had complete neuroimaging data, trait anxiety scores, blood- and physiological biomarker measurements. Trait Anxiety was assessed using the State-Trait Anxiety Inventory (STAI)[37, 38]. To capture perceived and chronic stress, we additionally included two validated instruments: the Perceived Stress Questionnaire (PSQ) [40] and the Trier Inventory for Chronic Stress (TICS) [41]. All preprocessed data were publicly available [102, 103]. The original LEMON dataset’s acquisition adhered to the Declaration of Helsinki principles and received approval from the University of Leipzig’s Medical Faculty Ethics Committee (reference number 154/13-ff) [102]. As this study exclusively utilized de-identified, publicly available data, no additional institutional ethical approval was required.

### 6.2 Neuroimaging Data

Neuroimaging data were acquired from the LEMON dataset, utilizing a 3 Tesla Siemens MAGNETOM Verio scanner equipped with a 32-channel head coil. We utilized resting-state blood oxygen level dependent (BOLD) time-series signals, diffusion MRI (DWI) and T1-weighted imaging for registration. For a detailed description of the MRI sequences employed for data acquisition, please refer to the original dataset publication [102].

All neuroimaging data were preprocessed by the original LEMON dataset researchers and are publicly available [102, 103]. Briefly, eyes-open rs-fMRI data underwent essential preprocessing steps including motion correction, distortion correction, denoising and band-pass filtering between 0.01-0.1 Hz, followed by spatial normalization to MNI152 space. To reduce boundary artifacts and non-stationarity, the first and last 10% of volumes were discarded and each ROI time series was z-scored (TR=1.4s, timepoints = 522). For DWI, the preprocessing included denoising, eddy current and susceptibility distortion correction and whole-brain probabilistic tractography. Structural connectivity was quantified as streamline counts, with fiber crossing and disconnection effects corrected via the SIFT2 method [104].

Preprocessed structural connectivity, functional connectivity and resting state fMRI time-series data are publicly available[103]. Briefly, brain regions were parcellated using data-driven atlases provided with the dataset. These atlases offer multiple parcellation scales, ranging from 183-2165 micro-regions, generated via voxel-wise clustering of rs-fMRI signals within macro-anatomical regions [103]. This data-driven approach aims to capture modular correspondences between structural and functional networks often missed by anatomical templates [105, 106]. For our primary analysis, we utilized the 183-region parcellation and the 391-region parcellation (Supplementary Materials) was employed for supplementary analyses to assess the robustness of our findings across different spatial resolutions.

#### 6.2.1 Amplitude of Low-Frequency Fluctuations (ALFF) computation

Amplitude of low-frequency fluctuations (ALFF) was computed from resting-state fMRI time series (TR = 1.4 s; sampling frequency *f*_*s*_ = 0.714 Hz derived from TR) extracted from 183 regions of interest (ROIs). Following established BOLD frequency partitions [107], ALFF was estimated separately for the *slow-5* (0.01–0.027 Hz) and *slow-4* (0.027–0.073 Hz) bands. A zero-phase 4th-order Butterworth band-pass filter was applied prior to spectral estimation to minimize phase distortion and spectral leakage. Power spectral density (PSD) was computed for each ROI using Welch’s method with a segment length of 256 time points and 50% overlap. The ALFF value for each ROI was defined as the area under the PSD curve within the corresponding frequency band, integrated using the trapezoidal rule, representing total oscillatory power in that band [108, 109]. This yielded a 183-dimensional ALFF vector per subject, which served as band-specific amplitude features for downstream analyses.

### 6.3 Allostatic Load Related Physiological Data

Allostatic load markers were assessed using blood- and physiological biomarkers derived from the dataset. These markers were selected for their established roles as mediators across cardiovascular, metabolic and immune systems, aligning with common practice in anxiety research [110–112]. Specifically, the included biomarkers were:

- **Cardiovascular:** Systolic Blood Pressure (SBP, mmHg) and Diastolic Blood Pressure (DBP, mmHg), measured at the left arm before the MRI session.
- **Metabolic:** Body Mass Index (BMI, kg/m^2^), Total Cholesterol (mmol/l), Low-Density Lipoprotein Cholesterol (LDL-C, mmol/l), High-Density Lipoprotein Cholesterol (HDL-C, mmol/l), Total Cholesterol to HDL-C Ratio and Glucose (mmol/l).
- **Immune & Renal:** Creatinine (*µ*mol/l) and C-Reactive Protein (CRP, mg/l).

Blood samples were collected approximately 70 ml in total, utilizing 4 different tubes (Serum, EDTA, Citrate, RNA) and processed according to the LEMON dataset’s standard procedure [102]. It was collected after of MRI data, however if it was not possible on the day it was acquired on the following assessment days. For our analysis, only data collected within 72 hours of the MRI acquisition were utilized to ensure the temporal consistency between neuroimaging and physiological markers. From an initial cohort of 132 participants with complete functional and structural neuroimaging data, 12 individuals were excluded due to either missing values for one or more of the selected allostatic load biomarkers or failing to meet the collection window relative to MRI acquisition, resulting a final sample of 120 participants for analysis.

### 6.4 Transcriptomic data

Transcriptomic features were obtained from the openly available preprocessing provided by Jimenez et al. (2024) [103], who processed the Allen Human Brain Atlas (AHBA) data [113] using the abagen toolbox[114] with default parameters. Full details of the preprocessing pipeline are available in (https://github.com/compneurobilbao/bha2).

### 6.5 Model Architecture

Our framework predicts trait anxiety by integrating brain and physiological data through three modules: a Graph Attention Network [115] for brain representation (NeuroGAT), an Allostatic Load (AL) Projection module for physiological representation and a Cross-Modal Attention module for information fusion, followed by a final regression head (**Figure 1a**). Unimodal baseline model architectures are detailed in the Supplementary Materials.

#### 6.5.1 Neuro Graph Attention Network (NeuroGAT)

The NeuroGAT models brain representation using rs-fMRI time series as node features and binarized structural connectivity as edges. To evaluate temporal and frequency information, we implemented four node feature extraction strategies:

- **Frequency-Filtered Multi-Layer Perceptron(MLP):** Per-band two-layer MLP with LeakyReLU was applied to slow-4 (0.027–0.073 Hz) and slow-5 (0.01–0.027 Hz) band-pass filtered rs-fMRI signals and the resulting per-ROI embeddings were combined via softmax-weighted concatenation.
- **Frequency-Filtered 1 Dimensional Convolutional Neural Network(1DCNN):** Per-band 1DCNN (kernel = 7) was applied to slow-4 and slow-5 band-pass filtered signals and per-ROI embeddings were combined via softmax-weighted concatenation.
- **Frequency-Agnostic MLP:** Two-layer MLP with LeakyReLU was applied to unfiltered rs-fMRI time series to produce per-ROI embeddings.
- **Frequency-Agnostic 1DCNN:** Single 1DCNN (kernel = 7) was applied to unfiltered rs-fMRI time series to produce per-ROI embeddings.

Three GATv2Conv layers with residual connections, each followed by LayerNorm, LeakyReLU and dropout (0.2). Node embeddings were pooled using Attentional Aggregation, yielding a 64-dimension global brain representation.

#### 6.5.2 Attention Weights

In the NeuroGAT, each edge (*i, j*) is assigned an attention coefficient *α*_*ij*_ that quantifies the relative contribution of node *j* to the representation of node *i*. Following the GATv2 formulation [116], attention scores are computed as

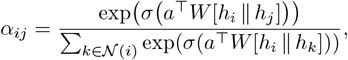

where *h*_*i*_ and *h*_*j*_ are node features, *W* is a learnable weight matrix, *a* is the attention vector and *σ* is the LeakyReLU nonlinearity. These coefficients are normalized across all neighbors of node *i* by a softmax function.

For model interpretation, we extracted *α*_*ij*_ from the trained model as edge-level attention values. Node-level importance was then defined as the mean of incident edge attentions. All reported attention maps represent averages across attention layers, outer cross-validation folds and repeated settings.

#### 6.5.3 Allostatic Load (AL) Projection

The AL Projection module transforms the ten allostatic load biomarkers into a dimensional representation. Prior to inputting into the module, all allostatic load parameters were standardized (z-score) for normalization within their respective training, validation and test sets. This module consists of two-layer MLP with a Leaky ReLU activation function applied after the first linear layer and dropout regulation (0.2). The AL representation is 64 dimensional output.

#### 6.5.4 Cross-Modal Attention

To fuse brain and physiological representations, we applied a two-head cross-modal attention. Formally, the brain embedding *h*_*brain*_ served as the query (*Q*) and the allostatic load embedding *h*_*AL*_ as both key (*K*) and value (*V*):

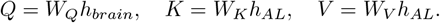

Attention was computed as

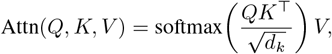

and outputs from two heads were concatenated and projected to 64 dimensions. A residual connection and layer normalization yielded the fused representation (64 dimensions).

Finally, the fused representation from the Cross-Modal Attention module is passed through a simple regression head to estimate the individual’s trait anxiety score. This head consists of two linear layers: an initial layer projecting to a 32 dimensional hidden layer with a Leaky ReLU activation function, followed by a final linear layer that outputs a single value representing the predicted trait anxiety score.

### 6.6 Training and Evaluation

All experiments were conducted in the Google Colab environment (Python 3.12.11, CUDA-enabled GPUs).

Models were trained and evaluated using a nested 3-fold cross-validation approach. This process was repeated 5 times, each with a different, arbitrarily chosen random seed to generate distinct train/validation/test splits. This strategy ensures robust and reproducible performance estimation across varied data partitions. The nested cross-validation involved an outer loop for unbiased performance evaluation and an inner loop for hyperparameter tuning. For specific performance comparisons or ablation studies, the same data splits were utilized.

Within each inner fold, hyperparameters were selected based on the mean *R*^2^ performance on the validation set. The hyperparameter search space included learning rates of 0.001, 0.0005, 0.0001 and 0.00005 and weight decay values of 0.01 and 0.001.

All models utilized the AdamW optimizer and Mean Squared Error (MSE) loss. A warm-up cosine decay learning rate scheduler was applied, with a 10-epoch warm-up phase followed by a cosine decay to a minimum learning rate of 1 *×* 10^*−*6^. Training proceeded for up to 300 epochs with a batch size of 32. To prevent overfitting, early stopping was implemented with a patience of 20 epochs and a dropout rate of 0.2. Final model performance was reported using averaged *R*^2^, MSE and Pearson correlation coefficient with actual trait anxiety score on the held-out test sets of five repeated cross-validated experiments.

#### 6.6.1 Comparison Models

Several comparison models were trained under identical conditions, including the same subject splits, five repeated nested 3-fold cross-validation and identical evaluation metrics.

The AlloNeuroGAT model with ALFF node features shared the same architecture and training configuration as the main full model, except that node features consisted of concatenated *slow-4* (0.027–0.073 Hz) and *slow-5* (0.01–0.027 Hz) ALFF values per ROI instead of temporally encoded features.

The Support Vector Machine (SVM) regressor was trained with the same data partitions, using as input each ROI’s ALFF values of *slow-4* and *slow-5* together with 10 physiological variables. Its hyperparameters were optimized using a grid search based on the negative mean absolute error (neg-MAE) from the inner validation folds, with the search space defined as *C ∈ {*0.1, 1, 10, 100*}, γ ∈ {*10^*−*4^, 10^*−*3^, 10^*−*2^, auto*}* and kernel *∈ {*rbf, linear*}*.

The XGBoost regressor was evaluated using the same cross-validation scheme and the same input features as the SVM model, with hyperparameters tuned over n_estimators = 180, max_depth *∈ {*3, 5, 7*}*, colsample_bytree *∈ {*0.7, 0.9, 1.0*}*, min_child_weight *∈ {*1, 5*}* and reg_lambda *∈ {*1.0, 5.0, 10.0*}*.

The Multilayer Perceptron (MLP) baseline used the same repeated nested cross-validation protocol and input features as the SVM and XGBoost models, with the AdamW optimizer, warm-up cosine decay scheduler, batch size of 32, maximum of 300 epochs and early stopping with a patience of 20, matching all main-experiment training settings.

### 6.7 Attention-Derived Statistical Analysis

All statistical analyses were conducted in Python (version 3.12.11) using the `statsmodels` package (version 0.14.0) and the `scipy` package (version 1.16.2) within the Google Colab environment.

#### 6.7.1 Mapping ROIs to Yeo 7 Networks

Each ROI was assigned to one of the Yeo 7 functional networks using the Yeo2011 7-network atlas (`MNI152_FreeSurferConformed1mm_LiberalMask.nii.gz`) ROI MNI centroids were converted to voxel indices and network labels were determined via local majority vote. Specifically, all voxels within a 1 mm spherical neighborhood around each centroid were extracted and the most frequent label within that sphere was assigned to the ROI. For ROIs with multiple coordinate entries, we assigned the mode of these local labels as the final network label for that entire ROI. Voxels outside the atlas bounds were coded as -1 (unassigned).

#### 6.7.2 Network-wise Attention Value

Edges were treated as undirected and labeled by the Yeo-7 network(s) of their incident nodes. Intra-network attention was the mean attention across edges with identical network labels; inter-network attention for a pair (*A, B*) was the mean across edges with endpoints in *A* and *B*. Heterogeneity across networks was tested with Kruskal–Wallis and FDR-BH–corrected Dunn tests; intra vs inter comparisons used Mann–Whitney *U* with FDR-BH across tested pairs.

#### 6.7.3 Consensus Community Detection

To examine whether attention patterns recapitulated functional systems, we applied consensus community detection on node-level importance derived by averaging attention across all incident edges. We used the Leiden algorithm (150 runs, ModularityVertexPartition, resolution=1.0, n_iterations=-1 until convergence). This matrix was clustered using agglomerative clustering across candidate community numbers (2–10) and the solution with the highest silhouette score (0.9182) was chosen as the final consensus partition.

#### 6.7.4 Mutual Information Correspondence

Correspondence between attention-derived communities and Yeo-7 labels was quantified with normalized mutual information. Significance was assessed via permutation testing by randomly shuffling community labels across ROIs (*N* =10,000 permutations).

### 6.8 Dynamic Functional Connectivity Analysis

#### 6.8.1 Dynamic functional connectivity estimation

Dynamic fluctuations in whole-brain functional connectivity (dFC) were estimated using a sliding-window correlation approach. For each participant, windowed FC matrices were computed with a 30-TR window and 3-TR step from bandpass-filtered BOLD signals (*slow-4*: 0.027–0.073 Hz; *slow-5*: 0.01–0.027 Hz). Windows showing local maxima in FC variance were selected as subject-specific exemplars to capture moments of maximal network reconfiguration [117].

##### State Identification

To identify and characterize recurrent dFC patterns, all exemplars from the entire sample were concatenated and clustered using spherical *k*-means with cosine similarity, which captures pattern similarity independent of magnitude [117]. The optimal number of clusters (*k* = 4) was determined using the elbow criterion and clustering was repeated 50 times with random centroid initializations to minimize local minima effects. Each participant’s complete set of dFC windows was then reassigned to the derived centroids, yielding a temporal sequence of discrete state labels representing distinct connectivity configurations.

##### Temporal and network-level metrics

For each state *s*, the dwell fraction *D*_*s*_ was computed to quantify how long each participant remained in that connectivity configuration:

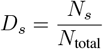

where *N*_*s*_ is the number of windows assigned to state *s* and *N*_total_ is the total number of windows for that participant.

To visualize and interpret inter-network organization, for each state, we averaged all participants’ FC windows assigned to that state to obtain a group-level mean FC matrix, which was then averaged within and between Yeo-7 functional networks to produce a network-level coupling map.

To visualize and quantify the network organization of each dFC state, we constructed thresholded graph representations based on the state-wise mean FC matrices based on prior literature [118]. For each state, the upper-triangular elements of the mean FC matrix (*C*_*k*_) were thresholded at the top 20% of positive correlations to retain only the strongest functional links. The resulting adjacency matrix was used to generate a graph with nodes corresponding to ROIs and edges representing supra-threshold connections. Only ROIs belonging to the Yeo-7 canonical networks were included to ensure functional interpretability. The graphs were visualized using the Fruchterman–Reingold force-directed layout implemented in `NetworkX`, with node colors denoting Yeo-7 network membership.

### 6.9 Permutation Feature Importance for Allostatic Markers

We estimated permutation-based feature importance for each allostatic-load biomarker on the outer-fold test sets. For each biomarker, we ran *N* = 1000 permutations within the test subjects of each outer fold (3-fold outer CV), repeated across five repetitions that defined distinct outer splits and model checkpoints. For a given permutation, we loaded the corresponding trained our best model (weights fixed), replaced the target biomarker column in the allostatic vectors with a subject-wise shuffled version confined to the current test set (other inputs unchanged) and recomputed predictions. Feature importance was defined as the degradation in test performance relative to the unpermuted baseline (primary metric Δ*R*^2^; ΔMSE and Δ*r* reported in Supplementary Materials). The permutation distributions were summarized by mean*±*SD.

### 6.10 Gene Ontology Analysis

Regional node attention scores were correlated with transcriptomic gradients from the Allen Human Brain Atlas (AHBA) and genes surviving Bonferroni correction (*p*_corrected_ *<* 0.05) were retained for enrichment analysis. Functional annotation was first conducted using the Metascape web platform (https://metascape.org), with both input and analysis species set to *H. sapiens* and the “Express Analysis” workflow applied. Metascape integrates multiple ontology and pathway resources; in our analysis, the significant gene set was evaluated across Gene Ontology (GO Biological Processes), DisGeNET disease enrichment, COVID-related pathways, PaGenBase (tissue-specific expression), Transcription Factor Targets and TRRUST regulatory networks. Non-redundant clusters were automatically generated by Metascape to summarize enriched terms.

To confirm the robustness of the molecular enrichment results, we performed an independent analysis using the same gene list with Enrichr [44]. Enrichment was assessed across multiple curated gene libraries, including KEGG, BioPlanet, Gene Ontology Biological Process (GO BP), Gene Ontology Cellular Component (GO CC) and Jensen Diseases (2025 Experimental).

## Acknowledgements

Nothing to Acknowledge.

## Data availability

All datasets analyzed in this study are publicly accessible. Raw MRI and behavioral data were obtained from the Leipzig Study for Mind–Body–Emotion Interactions (LEMON) dataset [102] (https://fcon_1000.projects.nitrc.org/indi/retro/MPI_LEMON.html). Transcriptomic data were sourced from the Allen Human Brain Atlas (https://human.brain-map.org/). Post-processed derivatives used in this work, including regional fMRI time-series, structural connectivity and functional connectivity matrices, are available at Zenodo (https://zenodo.org/record/8158914) [103]. No new data were generated in this study.

